# Integration of Expression QTLs with fine mapping via SuSiE

**DOI:** 10.1101/2023.10.03.23294486

**Authors:** Xiangyu Zhang, Wei Jiang, Hongyu Zhao

**Affiliations:** Department of Biostatistics, School of Public Health, Yale University, New Haven, Connecticut, United States of America

## Abstract

Genome-wide association studies (GWASs) have achieved remarkable success in associating thousands of genetic variants with complex traits. However, the presence of linkage disequilibrium (LD) makes it challenging to identify the causal variants. To address this critical gap from association to causation, many fine mapping methods have been proposed to assign well-calibrated probabilities of causality to candidate variants, taking into account the underlying LD pattern. In this manuscript, we introduce a statistical framework that incorporates expression quantitative trait locus (eQTL) information to fine mapping, built on the sum of single-effects (SuSiE) regression model. Our new method, SuSiE^2^, connects two SuSiE models, one for eQTL analysis and one for genetic fine mapping. This is achieved by first computing the posterior inclusion probabilities (PIPs) from an eQTL-based SuSiE model with the expression level of the candidate gene as the phenotype. These calculated PIPs are then utilized as prior inclusion probabilities for risk variants in another SuSiE model for the trait of interest. By leveraging eQTL information, SuSiE^2^ enhances the power of detecting causal SNPs while reducing false positives and the average size of credible sets by prioritizing functional variants within the candidate region. The advantages of SuSiE^2^ over SuSiE are demonstrated by simulations and an application to a single-cell epigenomic study for Alzheimer’s disease. We also demonstrate that eQTL information can be used by SuSiE^2^ to compensate for the power loss because of an inaccurate LD matrix.

**Author summary:** Genome-wide association studies (GWASs) have proven powerful in detecting genetic variants associated with complex traits. However, there are challenges in distinguishing the causal variants from other variants strongly correlated with them. To better identify causal SNPs, many fine mapping methods have been proposed to assign well-calibrated probabilities of causality to candidate variants. We introduce a statistical framework that incorporates expression quantitative trait locus (eQTL) information to fine mapping, which can improve the accuracy and efficiency of association studies by prioritizing functional variants within the risk genes before evaluating the causation. Our new fine mapping framework, SuSiE^2^, connects two sum of single-effects (SuSiE) models, one for eQTL analysis and one for genetic fine mapping. The posterior inclusion probabilities from an eQTL-based SuSiE model are utilized as prior inclusion probabilities for risk variants in another SuSiE model for the trait of interest. Through simulations and a real data analysis focused on Alzheimer’s disease, we demonstrate that SuSiE^2^ improves fine mapping results by simultaneously increasing statistical power, controlling the type I error rate, and reducing the average size of credible sets.

## Introduction

Over the past decades, genome-wide association studies (GWASs) have achieved remarkable success in detecting thousands of genetic variants that are associated with complex traits [1]. While GWASs have proven powerful in identifying genomic loci harboring causal variants, they encounter challenges in identifying the underlying causal variants. There is limited statistical power to distinguish causal variants from other variants in strong linkage disequilibrium (LD) through marginal association analysis [2, 3].

Genetic fine-mapping aims at inferring the causal genetic variants responsible for complex traits in a candidate region through disentangling LD patterns. Many fine mapping methods have been devised to assign well-calibrated probabilities of causality to candidate variants, taking into account the underlying LD pattern. For instance, some methods in the early stage estimate the probability of causality for each SNP under the assumption that each risk locus only harbors one causal variant [4, 5]. To avoid this strict assumption, CAVIAR [6] estimates the posterior inclusion probability (PIP) of each variant as a causal factor by jointly modeling the observed association statistics among all risk variants. Because of the heavy computational burden, CAVIAR makes the assumption that the total number of causal SNPs in a region is bounded by at most six, which leads to a major limitation. Under a similar statistical model, FINEMAP [7] enhances the computational efficiency by replacing the exhaustive search algorithm in CAVIAR with a shotgun stochastic search. However, this method is still computationally intensive. SuSiE [8], on the other hand, introduces a novel approach to variable selection in linear regression problems, where genetic fine-mapping is an important application. Building upon Bayesian variable selection in regression (BVSR), SuSiE develops an Iterative Bayesian Stepwise Selection (IBSS) algorithm to generate credible sets (CSs) that contain multiple highly correlated variables. The additive structure of the SuSiE model facilitates more accurate inference and improves computational efficiency, thereby enhancing the overall effectiveness of genetic fine-mapping.

In recent years, expression quantitative trait locus (eQTL) studies have revealed an abundance of quantitative trait loci (QTLs) for gene expression [9]. Integrating eQTL information into fine mapping not only improves the accuracy and efficiency of association studies by prioritizing functional variants within the risk genes but also aids in understanding the mechanisms underlying a genetic risk locus [10, 11]. Generally, there are two approaches to incorporating eQTL signals into fine mapping. The first approach involves conducting a colocalization analysis to determine whether the same variant is significant in both GWASs and eQTL studies. However, most colocalization methods, such as COLOC [12], eCAVIAR [13], and coloc-SuSiE [14], primarily focus on estimating the probability that a variant is causal in both GWASs and eQTL studies. This differs from our objective of identifying the causal variants associated with complex traits. The second approach incorporates gene expression levels as functional annotations and assigns functional priors to risk variants. Well established fine mapping methods incorporating annotations include PAINTOR [15], PolyFun+SuSiE [16], DAP [10], and SparsePro [17]. However, a significant drawback of the majority of these methods is that they are designed with two distinct modeling stages that employ different model settings for estimating prior probabilities and conducting fine mapping. This disjoint approach can result in potentially suboptimal performance [18].

In this study, we propose a new method of incorporating eQTL information to improve fine mapping results based on the SuSiE framework. Our new method, named SuSiE^2^, begins by prioritizing risk variants using estimated PIPs from an eQTL-based SuSiE model with expression levels of risk genes serving as the phenotype. These PIPs are then utilized as prior inclusion probabilities in a standard SuSiE model for the trait of interest. Through simulations conducted on UKBiobank samples, we demonstrate that compared with SuSiE, SuSiE^2^ improves the power of detecting causal SNPs while reducing false positives regardless of using the in-sample LD matrix or an external reference panel. For real data analysis, SuSiE^2^ identifies more Alzheimer’s disease (AD) associated SNPs predicted from single-cell epigenomic data.

## Materials and methods

### Posterior inclusion probabilities and credible sets

Consider a toy example of a multiple regression model between a standardized *n*-vector **y** and a standardized *n × p* matrix **X** = (*x*_1_, …, *x*_*p*_):

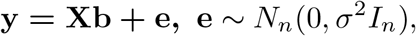

where **b** = (*b*_1_, …, *b*_*p*_)^*T*^ is a *p*-vector of regression coefficients, *σ*^2^ is the residual variance, *I*_*n*_ stands for the *n* × *n* identity matrix, and *N*_*n*_ represents the *n*-variate normal distribution. Many regression-based methods have been developed to select the associated variants, however, it can be difficult to infer the true causal variants when the effect variables are highly correlated with some non-effect variables (for example, genetic variants in strong LD). Under this circumstance, a more appropriate strategy is to narrow down a signal to a small set of highly correlated variants instead of an individual variant.

To quantify the uncertainty in which variables should be selected, BVSR methods [19] introduce a prior distribution on **b** and then calculate the posterior distribution that gives weights to each possible combination of causal variables. In most situations, this complicated posterior distribution is summarized with the marginal posterior inclusion probability (PIP) of each variable:

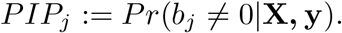

Although PIP provides a simple way to prioritize risk variants, it is somehow less informative and can be insufficient in determining true causal signals. For example, if the top two variants ranked by their PIPs are highly correlated, it is difficult to distinguish if they represent two different signals or if one of them is a non-effect variable correlated with a true causal one. With this consideration, a more appropriate result should provide a list of sets of variables, with each set intended to capture one signal. To describe this goal more formally, SuSiE (Sum of Single-effects regression model) [8] introduces the concept of a credible set of variables as below:

**Definition 1** *In a* multiple-regression model, a level *ρ credible set is defined to be a subset of variables that has probability ρ or greater of containing at least one effect variable*.

With this definition 1, the primary aim of the variable selection problem can be restated as the following two aspects:

1. Reporting as many credible sets as the data support, each with as few variables as possible.
2. Prioritizing each candidate variable within a credible set with a posterior probability for this variable to be an effect variable.

### The sum of single-effects regression model

With the goal of identifying the genetic variants that causally affect some traits of interest, genetic fine mapping can be framed as a variable-selection problem. To pick the causal variant(s) in the presence of strong LD, one attractive approach is to use BVSR to assign a posterior probability distribution to risk variants. However, traditional BVSR methods still suffer from computational challenges and complicated posterior distributions [7, 20], such as CAVIAR [6] and FINEMAP [7]. The SuSiE method introduced by [8] takes advantage of the convenient analytic properties of a more basic single-effect regression (SER) model [21] which only considers one effect variable with a non-zero regression coefficient. The SER model is described as:

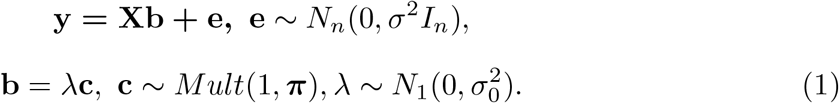

Here, **y** is the *n*-vector of the response variable, **X** = (*x*_1_, …, *x*_*p*_) is a matrix containing *n* observations of *p* explanatory variables, **b** is the *p*-vector of regression coefficients which can be decomposed as the product of a scalar *λ* and indicator variables **c** = (*c*_1_, …, *c*_*p*_)^*T*^ ∈ {0, 1}^*p*^, ***π*** = (*π*_1_, …, *π*_*p*_)^*T*^ gives the prior probability that each variable is the effect variable, *σ*^2^ and 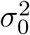 are the hyperparameters for the residual variance and prior variance of the non-zero effect. To avoid introducing an intercept term, **y** and the columns of **X** are assumed to have been centered to have zero means.

Under the SER model 1, there exists only one non-zero element in the coefficient vector **b**, determined by the indicator variables **c**. With fixed hyperparameters *σ*^2^ and 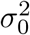, the posterior distribution of **b** = *λ***c** can be computed as:

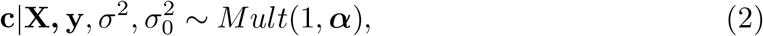

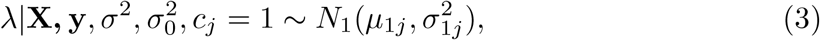

where ***α*** = (*α*_1_, …, *α*_*p*_)^*T*^ is the vector of PIPs, which can be computed with Bayes factors:

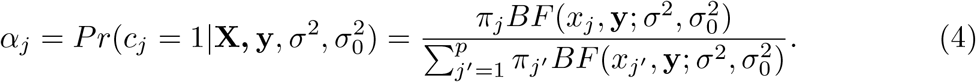

Here, *BF* (*x*, **y**; *σ*^2^, 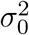) is the Bayes factor for comparing the following univariate linear regression model with the null model (*b* = 0):

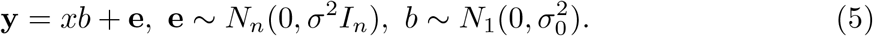

Suppose 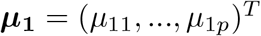, then the posterior distribution of **b** can be completely determined by ***α, μ***_**1**_, and 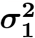, i.e., we can write the SER model as:

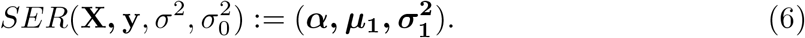

The SER model offers a simple inference strategy when there exists exactly one effect variable. However, the situation will be more complicated when there are multiple non-zero signals. To detect multiple effect variables while preserving the simplicity of the SER model, the sum of single-effects regression model (SuSiE) is developed by introducing multiple single-effect vectors and combining them with an additive structure:

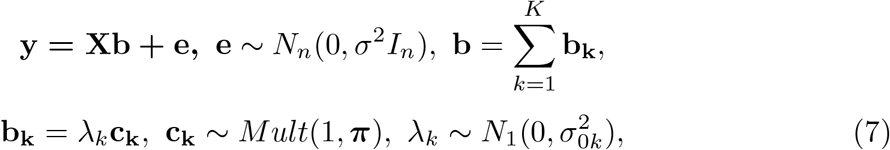

where **b**_**1**_, …, **b**_**K**_ represent the single-effect vectors each aiming to capture exactly one effect variable. 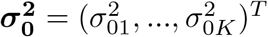 are the prior variances of the non-zero effects which can be different for different **b**_**k**_. *K* is the assumed total number of effect variables. A key feature of SuSiE is that, given **b**_**1**_, …, **b**_**K*−*1**_, estimating **b**_**K**_ simply involves fitting a SER model on residuals. This idea leads to the iterative Bayesian stepwise selection (IBSS) algorithm [8] in S1 Algorithm.

Different from existing BVSR models, SuSiE introduces a new model structure which naturally leads to an intuitive and fast algorithm for model fitting. Compared with traditional BVSR methods, SuSiE enjoys at least two key advantages:

1. SuSiE provides a posterior summary which can be interpreted easily by introducing the concept of “credible sets”.
2. SuSiE improves the computational efficiency, with a computational complexity *O*(*npK*). The running time of SuSiE, CAVIAR, and FINEMAP with the in-sample LD matrix and summary statistics has been compared in simulations [22], where SuSiE ran ten times faster than FINEMAP, and about 1,000 times faster than CAVIAR.

In the remaining parts of the method section, we will introduce a new framework to incorporate eQTL information into fine mapping based on SuSiE.

### Integrating eQTL information with fine mapping

Under the existence of strong LD, SuSiE assesses the uncertainty in variable selection by generating groups of variables, with each group aiming at capturing one effect variable. However, choosing the true causal variable from the credible set is still a difficult problem. One possible way to infer the effect variable more accurately is to integrate eQTL information into fine mapping, as SNPs associated with complex traits are significantly more likely to be eQTLs [11]. Considering the effect of each risk variable on the gene expression level helps us to prioritize risk SNPs with the posterior probability of being the effect variable, which can replace the prior distribution used in the original SuSiE manuscript: ***π*** = (1*/p*, …, 1*/p*)^*T*^.

This new framework of eQTL-based fine mapping study, named SuSiE^2^, connects two SuSiE models for eQTL study and genetic fine mapping, respectively. For the first model, we use the gene expression level as the response variable and conduct a regression analysis on the risk region. This eQTL-based SuSiE model can be rewritten as 8:

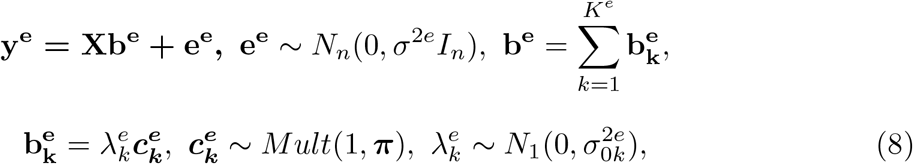

where **y**^**e**^ is the *n*-vector of gene expression levels, **b**^**e**^ is the *p*-vector of regression coefficients of risk variants for the gene expression, ***π*** is the naive prior inclusion probability for the eQTL-based SuSiE. Assume that there are in total *K*^*e*^ causal signals for the gene expression level, we can output from 8 the PIPs for all the single effects, denoted as 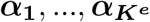. The final PIPs for the eQTL study can be computed as:

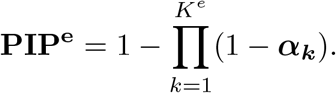

**PIP**^**e**^ represents the probability for each variant to be causal to the gene expression level. Under the assumption that trait-associated SNPs are more likely to be eQTLs, the PIPs from the eQTL-based SuSiE can serve as the prior distribution in the following SuSiE model for the trait of interest to highlight eQTLs in genetic fine mapping:

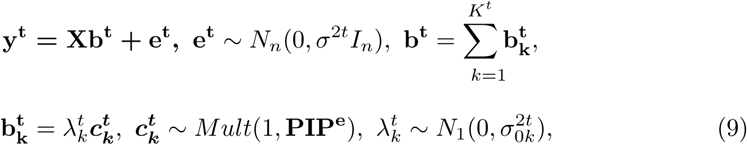

where **y**^**t**^ is the *n*-vector of trait of interest, **b**^**t**^ is the *p*-vector of regression coefficients of risk variants for this phenotype, and *K*^*t*^ is the total number of signals for the trait of interest.

Suppose from model 9 we detect single effects, with the corresponding PIPs denoted as 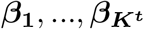, then the final PIPs for the trait of interest can be computed as:

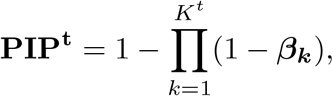

which prioritizes the candidate variants for the trait of interest. From model 9 we can also obtain the variants contained in credible sets for the trait of interest after adjusting for the eQTL priors.

In the method section above, we describe the SuSiE model and the SuSiE^2^ framework based on individual-level genotype data. We note that SuSiE has been extended to work with summary statistics [22], which makes it competitive with other well-developed fine mapping methods.

## Results

We demonstrate that integrating eQTL with fine mapping via SuSiE^2^ can indeed increase efficiency and accuracy through simulation studies and a real data study on Alzheimer’s Disease (AD). Compared with the original SuSiE, SuSiE^2^ can improve the results of fine mapping in the following aspects while controlling type I error rate at an appropriate level:

- SuSiE^2^ can improve the power of including causal variants in at least one credible set.
- SuSiE^2^ can decrease the average size for credible sets.

### Simulation

The study population in our simulations consists of 10,000 randomly selected Europeans from the UKBB dataset, with each sample genotyped at 20,000 SNPs on chromosome 1. Assuming a total of *L* risk loci associated with the trait of interest on this chromosome segment, we simulated the gene expression levels and the quantitative trait of interest through the following additive linear models:

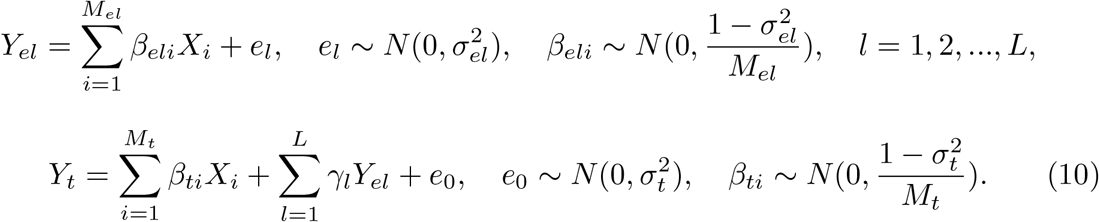

Here, *Y*_*el*_ is the gene expression level for the *l*th risk locus, *Y*_*t*_ is the quantitative trait of interest, *M*_*el*_ represents the number of causal SNPs for the *l*th risk locus, *M*_*t*_ is the number of causal SNPs for the trait of interest, *X*_*i*_ is the standardized genotype for the *i*th SNP, 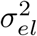 and 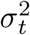 are the variance of error terms for the *i*th gene expression level and trait of interest, respectively. The effect sizes of the causal SNPs were assumed to follow normal distributions with zero means and variances chosen to ensure *V ar*(*Y*_*el*_) = *V ar*(*Y*_*t*_) = 1. For each risk locus, half of the *M*_*el*_ causal SNPs were also contained in the *M*_*t*_ effect variants for *Y*_*t*_. Therefore, the causal SNPs can affect the trait of interest either directly or through their effects on gene expressions, or in both ways.

The heritability for the trait of interest was selected from {0.1,0.2,0.3,0.4,0.5}, and the total number of causal SNPs was fixed at 30. These causal SNPs were equally distributed across *L* risk loci, with *L* being either 5 or 10. Throughout our simulations, we used the 95% percent credible sets to capture causal variants. We compared the performance of the following three methods: the original SuSiE without eQTL information (SuSiE), the SuSiE^2^ method that only used eQTL information from one risk locus (SuSiE2 partial), and SuSiE^2^ that used eQTL information from all the 5 or 10 risk loci (SuSiE2 all) with the following three criteria:

- Power: the proportion of true effect variables included in at least one credible set.
- Type I error rate: the proportion of non-causal variables included in at least one credible set.
- Average size: the average size of credible sets detected.

We first compared the results with summary statistics and the in-sample LD matrix, with the results summarized in Figure 1. We observed that for every combination of true heritability and number of risk regions, two SuSiE^2^ methods always improved the power of detecting causal SNPs and also reduced the average size of credible sets, and the improvement of SuSiE2 all was more significant compared with SuSiE2 partial. All three fine mapping approaches controlled the type I error rate at a low level with the 95% credible sets, but SuSiE2 all always had fewer false positives. The credible sets we obtained from SuSiE were designed to contain at least one effect variable with 95% probability. However, the target type I error rate here was the proportion of non-causal variables incorrectly detected, which can be largely influenced by the average size and total number of credible sets. This explains why the type I error rate seems to be way lower than the nominal level. When the number of risk loci increased from 5 to 10, the performance of SuSiE^2^ regarding all the criteria improved more significantly. With 10 risk loci, SuSiE2 all improved the power of detecting causal SNPs by 10% while reducing false positives by 50% when utilizing the in-sample LD matrix. Besides, we observed a 40% reduction in the average size of credible sets obtained by SuSiE2 all compared with the original SuSiE. It is worth mentioning that although not as good as SuSiE2 all, SuSiE2 partial achieved better performance compared with the original SuSiE, which indicated that considering only a small proportion of eQTL information can still help improve the results of fine mapping.

**Fig 1.**
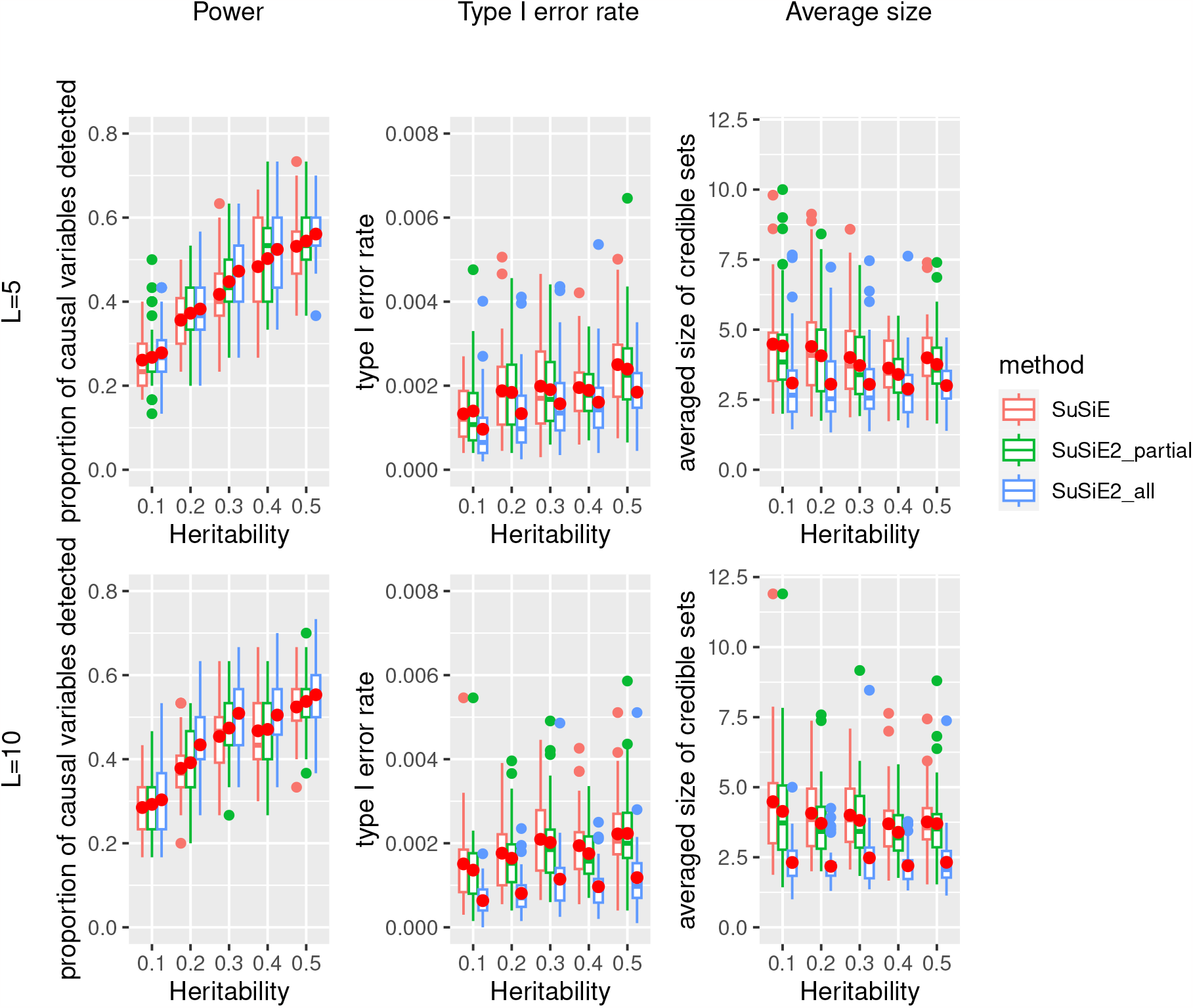
Simulation results of power, type I error rate, and averaged size of credible sets for three fine mapping methods with the in-sample LD matrix. This simulation was based on 10,000 UKBB samples and 20,000 SNPs with an in-sample LD matrix. The solid red dots represent the average values across 40 repetitions.

We also checked the performance of three fine mapping methods when using the LD matrix calculated from an external reference panel of 5,000 Europeans from the UKBB study, with the results summarized in Figure 2. All three methods controlled the type I error rate at a low level. The power of fine mapping studies based on the external reference panel was reduced on average and became less stable compared to the simulation using the in-sample LD matrix. This suggests that accurate information about the correlations between variants plays an important role in identifying the true causal variants. However, integrating the eQTL priors via SuSiE^2^ improved the performance of fine mapping for all situations. When the number of risk loci was 10, SuSiE2 all achieved better performance with a 50% increase in power and a 30% reduction in the proportion of false positives. This suggests that eQTL information can compensate for the power lost because of inaccurate LD information.

**Fig 2.**
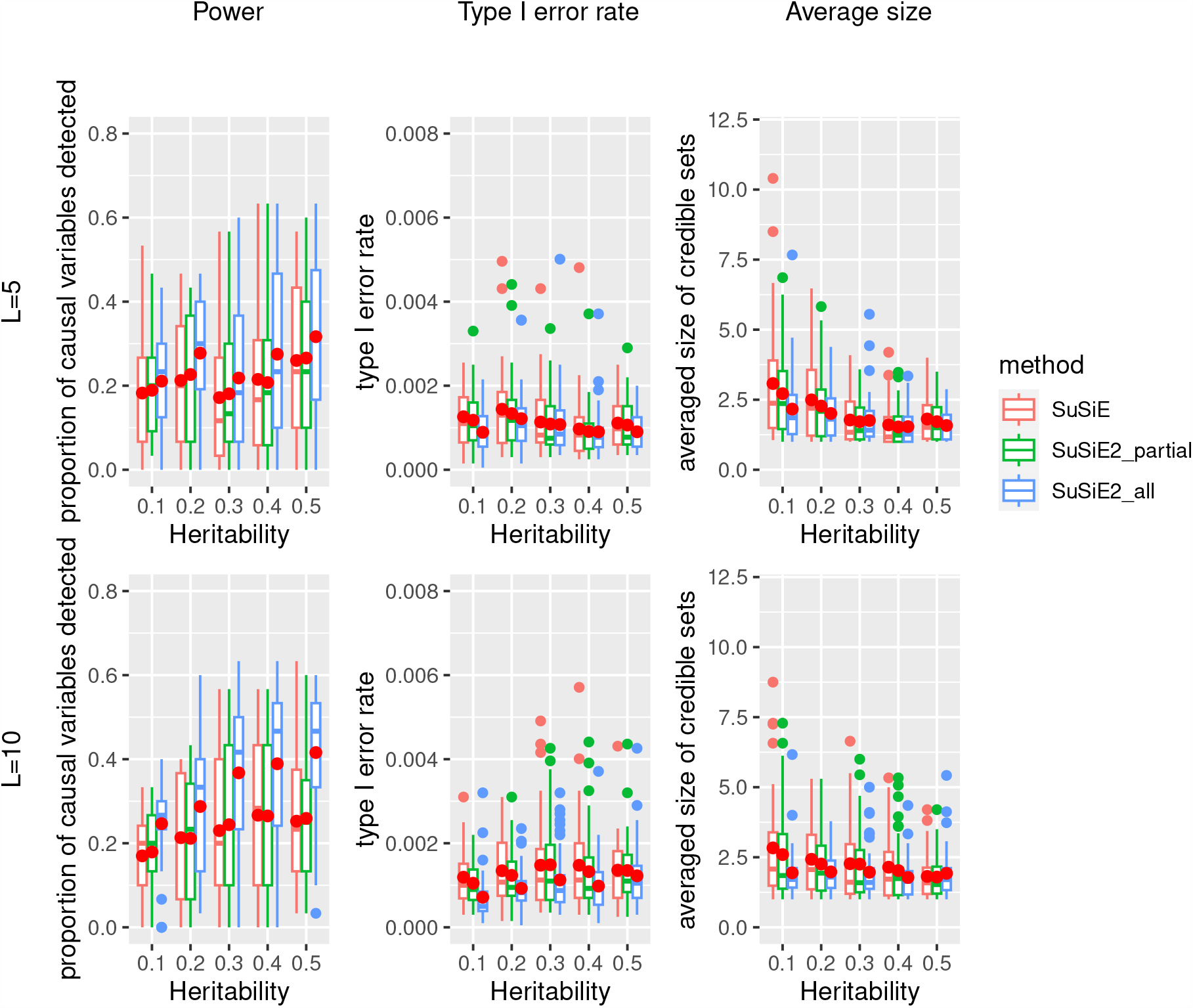
Simulation results of power, type I error rate, and averaged size of credible sets for three fine mapping methods with an external reference panel. This simulation was based on 10,000 UKBB samples and 20,000 SNPs with an external reference panel of 5,000 Europeans from the UKBB study. The solid red dots represent the average values across 40 repetitions.

### Application to AD dataset

In this section, we applied SuSiE^2^ to a real dataset on Alzheimer’s disease. The summary statistics we used were from a recent meta-analysis of individuals from 13 cohorts, with a total of 1,126,563 individuals (90,338 cases and 1,036,225 controls) included [23]. This meta-analysis identified 3,915 significant (*P <* 10^*−*8^) variants across 38 independent loci, including seven loci that had not been reported previously. The sample size generating the summary statistic for each SNP ranged from 216 to 762,917, with a median of 661,401. To make the z-scores of each SNP more comparable, we removed those SNPs with corresponding sample sizes smaller than 500,000.

We obtained the gene expression levels for AD risk loci from the ROSMAP dataset [24], which contained the bulk RNA sequencing (RNA-seq) data of 642 individuals. Among them, 473 individuals also had genotype data available on 572,266 SNPs, which allowed us to conduct an eQTL study for AD risk loci via SuSiE. We used the Michigan imputation server [25] with 1000 Genomes Phase 3 (Version 5) as the reference panel. After imputation, we obtained the genotype data for 473 ROSMAP samples at 13,753,668 SNPs.

To evaluate our method, we treated the predicted functional SNPs for Alzheimer’s diseases from a single-cell epigenomic analysis [26] as the validation data. This study developed a machine-learning classifier to integrate a multi-omic framework and identified multiple pairs of AD risk locus and the most likely mediator in both coding and non-coding regions. After removing the APOE locus because of multiple mediators, there were in total 35 pairs of AD risk locus and mediator, 16 in the coding regions and 19 in the non-coding regions.

Our real data analysis was conducted with the following steps:

1. We extracted all the common SNPs within 100kb upstream and downstream of each likely mediator as a target set.
2. The LD matrix was calculated for each target set with a reference panel based on Europeans from the UKBB dataset.
3. We fitted the eQTL-based SuSiE model with the ROSMAP dataset and calculated the PIP for each candidate SNP in the target set.
4. PIPs from step 3 were treated as prior distributions and integrated into the fine mapping study based on summary statistics from the meta-analysis to get SuSiE^2^ results.
5. Two fine mapping methods we considered were SuSiE^2^ and the original SuSiE that did not take advantage of the eQTL information. We only considered 20 mediator-risk loci pairs in the common part of the ROSMAP dataset, reference panel, and the meta-analysis dataset. We compared the AD mediators identified by SuSiE and SuSiE^2^, with the results summarized in Table 1. SuSiE^2^ successfully identified nine out of 20 mediators, while SuSiE only captured five of them. In the coding region, there were in total seven causal SNPs, SuSiE identified two of them, while SuSiE^2^ detected three of them. In the non-coding region, the number of AD mediators identified by SuSiE was three, while the number of mediators identified by SuSiE^2^ was six. We also evaluated the properties of generated credible sets (CSs) by two methods, summarized in Table 2. The original SuSiE captured 27 credible sets, with an average size of 9.6, while integrating eQTL information allowed us to identify 29 credible sets and reduced the average size to 8.0. Compared with SuSiE, SuSiE^2^ also reduced the 75% quantile of the size of credible sets from 13 to 11, which suggests that SuSiE^2^ may avoid producing extremely large credible sets.

**Table 1.** Summary of AD mediators detected by SuSiE and SuSiE^2^.

| Method | Total | Identified (Total) | Coding Region | Identified (Coding region) |
| --- | --- | --- | --- | --- |
| SuSiE | 20 | 5 | 7 | 2 |
| SuSiE<sup>2</sup> | 20 | 9 | 7 | 3 |

**Table 2.** Summary of credible sets identified by SuSiE and SuSiE^2^.

| Method | Number of CS | Average Size | 25% Quantile | Median | 75% Quantile |
| --- | --- | --- | --- | --- | --- |
| SuSiE | 27 | 9.6 | 2 | 4 | 13 |
| SuSiE<sup>2</sup> | 29 | 8.0 | 2 | 4 | 11 |

We also calculated the PIP for each mediator by SuSiE and SuSiE^2^, as shown in Figure 3. From this plot, we observed that SuSiE^2^ can identify more AD mediators by increasing the estimated PIPs of them, and all the mediators identified by SuSiE were also captured by SuSiE^2^. Besides, the points of many causal SNPs were distributed around the *y* = *x* line, which suggests that the SuSiE regression model may not be very sensitive to the choice of prior probabilities. The numerical results of PIPs estimated by SuSiE and SuSiE^2^ for every AD mediator are summarized in S1 Table.

**Fig 3.**
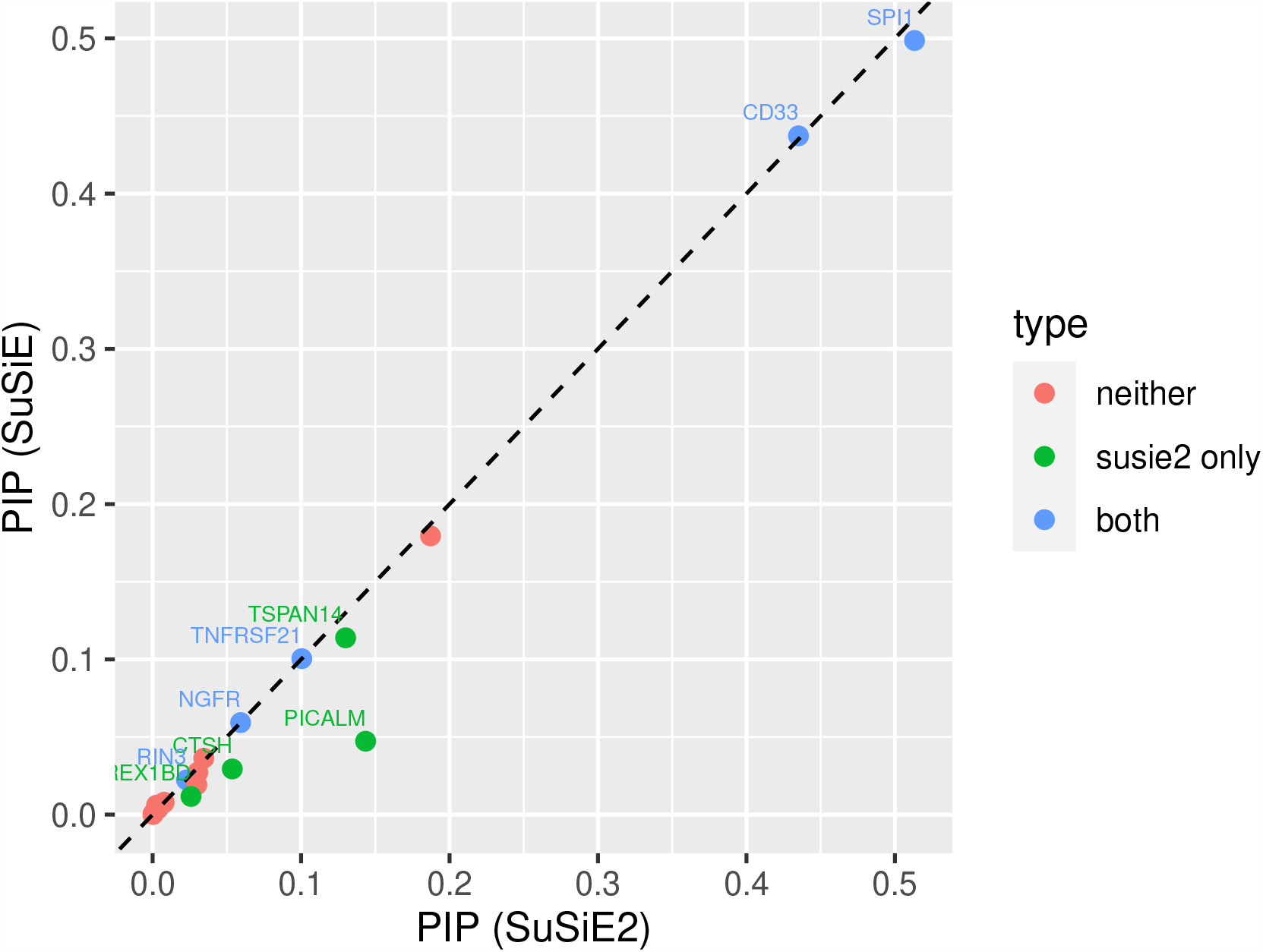
Estimated PIP for each AD mediator by SuSiE and SuSiE^2^. There were in total 20 AD risk loci divided into the following three categories. Five mediators were detected by both SuSiE and SuSiE^2^, denoted by the blue dots. SuSiE^2^ identified four additional risk loci, denoted by the green dots. The remaining 11 loci could not be detected by either SuSiE or SuSiE^2^, corresponding to the red dots.

To illustrate that SuSiE^2^ enhanced the PIPs for causal mediators, we display the examples of two risk loci in Figure 4. We considered the PIPs for all variants within these loci from the following three categories: eQTL study, SuSiE, and SuSiE^2^. The PIPs estimated from the eQTL study are used as the prior information by SuSiE or SuSiE^2^. For the PICALM locus (Figure 4 A), a slightly larger PIP was assigned to the true AD mediator compared with most candidate variants by the eQTL-based SuSiE, which allowed SuSiE^2^ to capture this mediator in a credible set. However, the original SuSiE failed to include this variant in any credible sets. For the C14orf93 locus (Figure 4 B), both SuSiE and SuSiE^2^ failed to find any signal in the risk locus. The estimated PIPs by SuSiE were stable at a very low level, with the largest PIP smaller than 0.05. In contrast, with the prior information provided by the eQTL study, the signals for some candidate SNPs in this region were enhanced with the strongest PIP larger than 0.15. Besides, the PIPs for the remaining SNPs estimated by SuSiE^2^ were reduced towards zero, which indicated that SuSiE^2^ performed better in separating causal SNPs from non-causal variants.

**Fig 4.**
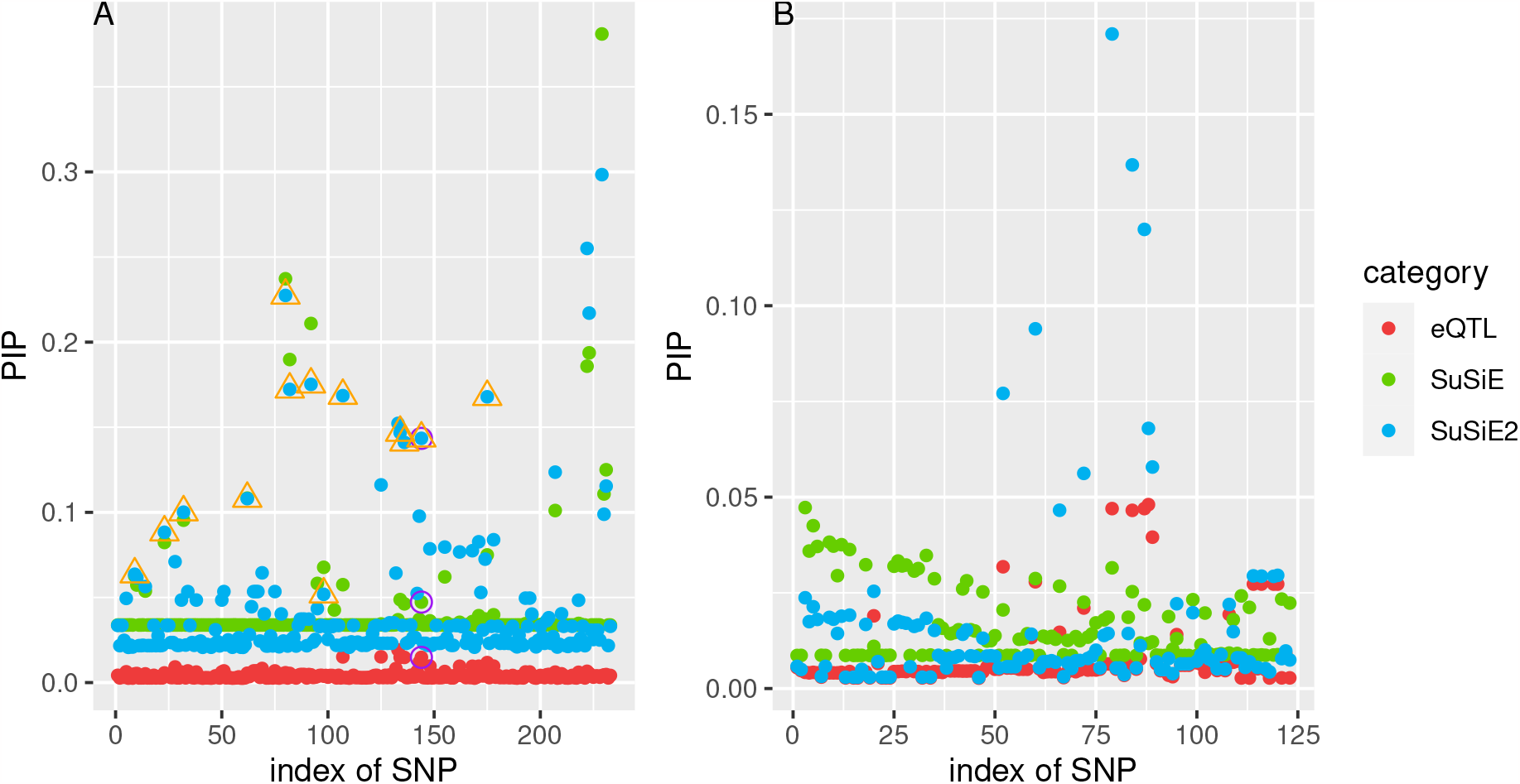
Estimated PIPs by SuSiE, SuSiE^2^ and eQTL-based SuSiE for PICALM (A) and C14orf93 (B). The PIPs estimated from the eQTL study are used as the prior information by SuSiE or SuSiE^2^. For the PICALM locus, PIPs for the true mediator in this locus are surrounded by the purple circle, and the points surrounded by an orange triangle correspond to the credible set from SuSiE^2^ which can capture the true mediator. For the C14orf93 locus, the true mediator was not included in the common part of summary statistics and ROSMAP data.

In conclusion, the real data analysis results on the AD dataset also suggest that incorporating eQTL information in the SuSiE model increased the statistical power of identifying the true variants while reducing the average size of credible sets. Besides, SuSiE^2^ achieved a better performance in separating causal SNPs from non-causal SNPs.

## Discussion

Statistical fine mapping has been an important tool in detecting the true causal SNPs for complex traits of interest. Most widely used fine mapping methods are based on the Bayesian framework, and assigning a proper prior distribution to risk variants can improve both the accuracy and efficiency of fine mapping. As an important indicator of association with gene expression level, eQTL information can be incorporated into fine mapping by either conducting a colocalization study or fine mapping with annotations. In this manuscript, we proposed a new framework for integrating eQTL with fine mapping via the SuSiE model. Through the simulation study, we showed that this new framework can increase statistical power while reducing the average size of credible sets. The advantage of SuSiE^2^ compared with the original SuSiE in improving the statistical power was more apparent when we used an external reference panel. The real data application in AD also suggests that SuSiE^2^ performed better than other methods in identifying the true AD mediators by prioritizing risk variants based on eQTL information before conducting the association study.

A number of issues remain to be addressed in the future. The first one is that the formulation of SuSiE^2^ may be improved so that we do not have to run SuSiE two times. In other words, we may accomplish the eQTL-adjusted SuSiE within one framework. Second, although our simulation suggests that SuSiE is generally robust to overstating of the total number of causal effects *K* in the IBSS algorithm [8], SuSiE was not very stable to the choice of *K* in real data applications. A larger *K* sometimes leads to the finding of new credible sets. Based on our experience, we recommend increasing the parameter *K* starting from 1 and stopping this process when we fail to find new credible sets. Further investigation of the mechanisms underlying this phenomenon is needed to find the best way to select the parameter and make use of the prior information. Third, as eQTLs may be context and cell-type specific, we may jointly consider eQTLs across multiple conditions and also include other molecular QTL information to more comprehensively capture different mechanisms contributing to diseases.

## Conclusion

In this manuscript, we have introduced SuSiE^2^, a statistical framework that incorporates eQTL information to fine mapping. By prioritizing variants within the candidate region with eQTL information, SuSiE^2^ improves the performance of fine mapping by simultaneously increasing statistical power, reducing false positives, and decreasing the average size of credible sets compared with the original SuSiE. We also demonstrate through simulations that eQTL information can compensate for the power loss because of inaccurate LD information. In the real data application, SuSiE^2^ confirms four more functional SNPs associated with AD predicted from single-cell epigenomic data compared with SuSiE. Evaluations of AD risk genes like PICALM and C14orf93 indicate that SuSiE^2^ enhances the PIPs for causal mediators and achieves superior performance in distinguishing causal SNPs from non-causal variants.

## Data Availability

We used the UK Biobank genotype data (https://www.ukbiobank.ac.uk/) and conducted the research using the UKBB resource under approved data request (access ref: 29900).
Moreover, the following data used in the real data analysis are publicly available:
The Alzheimer's Disease summary statistics: https://ctg.cncr.nl/software/summary_statistics
ROSMAP Gene Expression data: https://www.synapse.org/#!Synapse:syn17008934
The validation data of functional SNPs for AD was available in Supplementary Table 2 from https://doi.org/10.1038/s41588-020-00721-x

## Supporting information

**S1 Algorithm. Iterative Bayesian stepwise selection (IBSS) algorithm [8]**.

### Algorithm 1 IBSS

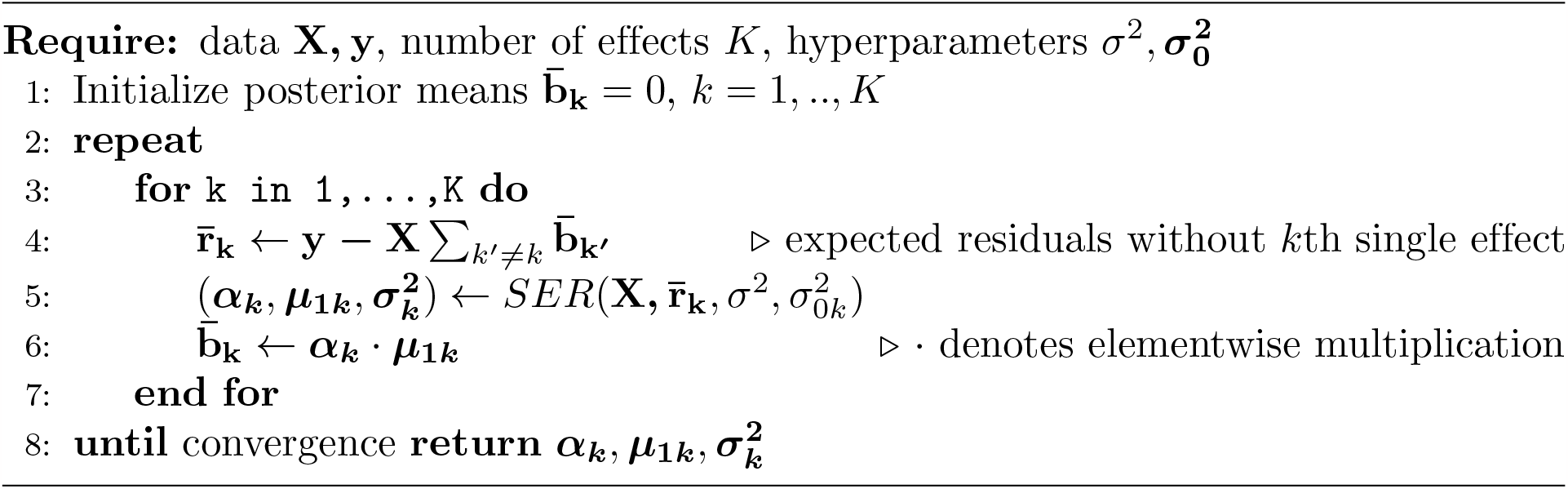

**S1 Table.** Summary information of AD mediators. We summarize the chromosome, SNP ID, AD risk gene, indicator of the coding region, whether or not this mediator can be identified by SuSiE and SuSiE^2^, and the estimated PIPs for every AD mediator in this table.

| Chromosome | SNP ID | Gene | Region | SuSiE | SuSiE<sup>2</sup> | PIP(SuSiE) | PIP(SuSiE<sup>2</sup>) |
| --- | --- | --- | --- | --- | --- | --- | --- |
| 1 | rs4575098 | ADAMTS4 | non-coding | FALSE | FALSE | 0.17953 | 0.18719 |
| 2 | rs13025717 | BIN1 | non-coding | FALSE | FALSE | 0.01919 | 0.02976 |
| 6 | rs1004173 | TNFRSF21 | non-coding | TRUE | TRUE | 0.10046 | 0.10046 |
| 7 | rs6464547 | TMEM139 | non-coding | FALSE | FALSE | 0.00743 | 0.00743 |
| 10 | rs7920721 | USP6NL | non-coding | FALSE | FALSE | 0.00006 | 0.00020 |
| 10 | rs7900536 | TSPAN14 | non-coding | FALSE | TRUE | 0.11388 | 0.13001 |
| 11 | rs2276412 | SORL1 | coding | FALSE | FALSE | 0.00612 | 0.00263 |
| 11 | rs3740688 | SPI1 | coding | TRUE | TRUE | 0.49861 | 0.51323 |
| 11 | rs1237999 | PICALM | non-coding | FALSE | TRUE | 0.04728 | 0.14347 |
| 14 | rs3829409 | C14orf93 | coding | FALSE | FALSE | 0.03634 | 0.03447 |
| 14 | rs10130373 | RIN3 | non-coding | TRUE | TRUE | 0.02232 | 0.02281 |
| 15 | rs2289702 | CTSH | coding | FALSE | TRUE | 0.02941 | 0.05366 |
| 15 | rs653765 | ADAM10 | non-coding | FALSE | FALSE | 0.00085 | 0.00048 |
| 15 | rs72749561 | MEX3B | non-coding | FALSE | FALSE | 0.00368 | 0.00361 |
| 17 | rs3816913 | USP6 | coding | FALSE | FALSE | 0.02734 | 0.03049 |
| 17 | rs28618326 | NGFR | non-coding | TRUE | TRUE | 0.05928 | 0.05928 |
| 19 | rs3764645 | ABCA7 | coding | FALSE | FALSE | 0.01769 | 0.02767 |
| 19 | rs12459419 | CD33 | coding | TRUE | TRUE | 0.43724 | 0.43503 |
| 19 | rs2303696 | REX1BD | non-coding | FALSE | TRUE | 0.01170 | 0.02585 |
| 20 | rs17462136 | CASS4 | non-coding | FALSE | FALSE | 0.00798 | 0.00777 |

The SuSiE column represents whether or not the original SuSiE can identify the corresponding AD mediator. The SuSiE<sup>2</sup> column is similar.

## Acknowledgments

This work was supported in part by the National Institutes of Health [R01 GM134005, U24 HG012108] and the National Science Foundation grant [DMS1902903]. We thank the participants of the UK Biobank and conducted the research using the UKBB resource under approved data request (access ref: 29900).

## Notes

### Competing Interest Statement

The authors have declared no competing interest.

### Author Declarations

The North West Multi-Centre Research Ethics Committee (MREC) of UK Biobank gave ethical approval for this work. (ref: 29900)

### Summary of Updates

Ref number corrected.

